# NeuroEPO improves cognition in Parkinson’s disease. Preliminary report

**DOI:** 10.1101/2022.02.24.22271444

**Authors:** Maria L. Bringas Vega, Liu Shengnan, Yanetsy Rodriguez Leon, Ania Mesa Rodriguez, Enrique Casabona Fernandez, Marite Garcia Llano, Daniel Amaro Gonzalez, Teresita Rodriguez Obaya, Leslie Perez Ruiz, Iliana Sosa Teste, Fuleah A. Razzaq, Marjan Jahanshahi, Ivonne Pedroso Ibañez, Pedro Valdes-Sosa

## Abstract

**Background:** Cognitive impairment is a feature of Parkinson’s Disease (PD) from the early stages, but currently, no treatment for cognitive deficits in PD is available. Erythropoietin (EPO) has been studied for its potential neuroprotective properties in neurologic disorders with a beneficial action on cognition.

**Objective:** To evaluate if NeuroEPO, a new formulation of EPO with low content of sialic acid, improves the cognitive function in PD patients.

**Methods:** A double-blind, randomized, placebo-controlled physician lead trial was conducted. The sample was composed of 26 PD patients (HY stages I-II), where 15 received intranasal NeuroEPO for 5 weeks, and another age and gender-matched 11 patients were randomly assigned to the Placebo. All the samples received 9 months of intensive NeuroEPO treatment during a post-trial. The cognitive functions were assessed using a comprehensive neuropsychological battery before, one week, and 6 months after the first intervention and one week after a 9months post-trial. The effects of NeuroEPO were evaluated using a multivariate linear mixed-effects model using a latent variable for cognition instead of the raw neuropsychological scores.

**Results:** A significant and direct effect of the Dose of NeuroEPO (p=0.00003) was found on cognitive performance with a strong positive influence of educational level (p=0.0032) and negative impact of age (p=0.0063).

**Conclusions:** These preliminary results showed a positive effect of NeuroEPO on cognition in PD patients with better benefit for younger and higher educated patients.

## Introduction

Parkinson’s disease (PD) is a neurodegenerative disorder that mainly affects the motor system, but with a range of non-motor symptoms, including cognitive dysfunction, depression, and anxiety ^1^. In the early stages, executive dysfunction ^2^ and other forms of mild cognitive impairment ^3^ are features of PD. In contrast, in the later stages of PD a significant number of patients develop dementia ^4^. Cognitive impairment highly impacts the quality of life in PD^5,6^. Currently, the motor symptoms of PD can be well controlled initially by dopaminergic medication ^7^ and in the later stages with deep brain stimulation ^8^, but there is no treatment for cognitive deficits in PD.

Erythropoietin (EPO) is a cytokine known as an essential hematopoietic growth factor in tissue oxygenation ^9^. It is a glycoprotein hormone with 165 amino acids, weighs 30.4 kDa, and is a member of the cytokine superfamilies. At present, it is widely used in the treatment of anemia related to premature births, renal failure, cancer, chronic inflammatory diseases, and HIV infections ^10^. EPO is believed to have functions that keep tissue oxygenation at adequate levels because its hematopoietic function^11^. Some evidence also established that EPO has other functions, such as neuroprotection, although the mechanisms are not fully clarified. Preclinical studies in PD and other neurological and psychiatric disorders have suggested the neuroprotection capacity of EPO ^12^.

But the treatment of neurological diseases with EPOrh involves a higher dose and prolonged application, producing adverse effects because of the increment of hematocrit and blood viscosity. For that reason, here we tested the impact of NeuroEPO, a new formulation of EPO with low content of sialic acid (molecule is between 4 and 7 mmol/mL of protein). This molecule is similar to that produced in the brain of mammals but does not have an inducer effect in the synthesis of erythrocytes, maintaining its neuroprotective properties.

NeuroEPO is safe and tolerated in healthy people^13^ and PD patients^14^, but its neuroprotective effects have been mainly tested in animal models ^15,16^, in-vitro models^17^, and partially reported ^18^ on the cognitive performance of PD patients. But this is the first attempt to study the long-term effects of cognitive effects of NeuroEPO in PD. We consider the importance of neuroprotection against the cognitive decline since 80% of PD patients in late-stage progress to dementia ^19^ and even in early stages, more than 25% have cognitive deficits ^20^.

The study aimed to assess the effect of NeuroEPO on cognitive function in PD patients in two trials, the 6-months randomized placebo-controlled safety trial where the participants received a small dose and later the long-term effect of the drug after a 9 month follow-up post-trial using a higher dose of NeuroEPO for all the participants.

To evaluate the effect of NeuroEPO, we employed the doses of the product received across the two trials; such a continuous process could be evaluated using a latent variables longitudinal linear dynamic model.

## Methods

The study was part of a randomized, double-blind physician-led placebo-controlled trial to evaluate the safety of NeuroEPO in Parkinson’s Disease patients (https://clinicaltrials.gov/ number NCT04110678) with motor and cognitive secondary outcomes measures. It was developed in collaboration between three institutions: the International Center for Neurological Restoration (CIREN) and the Center for Molecular Immunology (CIM), La Habana Cuba, and The Clinical Hospital of Chengdu Brain Sciences Institute, UESTC Chengdu, China.

The sample was composed of twenty-six patients (10 women) with a clinical diagnosis of idiopathic Parkinson’s disease according to the UK Brain Bank Criteria. The average age was 53.88 years (SD=7.66), and the duration of disease was 5.5 years (SD=3.49). The patients were in stages I-II of Hoehn and Yahr ^21^. The inclusion criteria were similar to those employed by our group in this safety trial^14^ and to test rHuEPO in another PD patients group^22^.

NeuroEPO [CIMAB S.A., Havana, Cuba] was a stabilized liquid formulation in a unique dose bulb containing 1 mg (1 mg/mL) of non-hematopoietic rHu-EPO, produced in Chinese hamster ovary (CHO) cells. Each bulb also contains buffer salts, polysorbate 80, sodium EDTA, NaCl, HPMC F4M, and water for injection to complete 1 mL.

The NeuroEPO group included fifteen patients (7 women) who received a weekly 1mL dose of intranasal NeuroEPO for 5 weeks, and the placebo group included eleven patients (3 women) who received a similar formulation containing the same ingredients except for EPO. Patients were randomly allocated to NeuroEPO or Placebo, and the two groups were matched for age and gender.

After the safety trial finished and following the therapeutic obligation principle (https://www.wma.net/policies-post/wma-declaration-of-helsinki-ethical-principles-for-medical-research-involving-human-subjects/) inherent to double-blind placebo-controlled trials, the sponsors and researchers conducted a post-trial where they gave access to NeuroEPO to all the patients of the safety trial. This information was disclosed to participants during the informed consent process.

All the patients (n=26) received for 9 months an intensive NeuroEPO treatment of 1mL bulb, 3 times per week, for one month (induction phase) and later 1/2 bulb three times per week, for eight months, summarizing a total of 60ml in all the periods in comparison with the 5 ml of the safety trial. Even if all the patients have taken part in the post-trial, only a subsample of 18 patients (10 from the original NeuroEPO group and eight from Placebo) attended the final session neuropsychological assessment nine months after the post-trial intervention. See **Table 1** for demographic and clinical data for the samples of both trials.

**Table 1.** Demographic and Clinical description of the sample.

| **Table 1. Demographic and Clinical description of the sample** |  |  |  |  |
| --- | --- | --- | --- | --- |
|  | **Group A** | **Group B** | **Total** | **Post-trial** |
|  | **NeuroEPO** | **Placebo** |  |  |
| **Total** | N=15 | N=11 | N=26 | N=18 |
| **Age [mean, SD]** | 56.4 ± 7.8 | 61.09 ± 6.6 | 58.4 ± 7.6 | 57 ± 11.9 |
| **Sex** |  |  |  |  |
| **Male** | 7 [46.6 %] | 8 [72.7%] | 15 [55%] | 12 [66.6%] |
| **Female** | 8 [53.4%] | 3 [27.2%] | 11 [45%] | 6 [33.3%] |
| **Stage Hoehn & Yahr** |  |  |  |  |
| **I** | 4 [26.6%] | 1 [9.09%] | 5 [19.2%] | 3 [16.7%] |
| **II** | 11 [73.4%] | 10 [90.9%] | 21 [80.8%] | 15 [83.3%] |
| **Duration of illness (years)** |  |  |  |  |
|  | 5.4 ± 3.2 | 5.8 ± 4.06 | 5.6 ± 3.5 | 5.1 ± 4.2 |
| **PD Familial antecedents** |  |  |  |  |
| **Yes** | 6 [40%] | 3 [27.2%] | 9 [34.6%] | 6 [33.3%] |
| **No** | 9 [60%] | 8 [72.8%] | 17 [65.4%] | 12 [66.6%] |
| **Levodopa equivalent Dose LED** | 935.83 ± 302.54 | 939.3 ± 194.0 |  |  |

The Dose for each participant is the cumulative amount of drug taken at the time of evaluation: 0 at the baseline for all the patients, 5 mL for NeuroEPO group, and 0mL for the Placebo group in the timepoint 1 (first week after complete intervention) and time point 2 (six months) assessments after first intervention respectively. During the post-trial, all the patients received 60 mg, a cumulative 60 mg for the original placebo group, and 65 for the NeuroEPO. See **Figure 1** for a detailed description of the trial.

**Figure 1.**
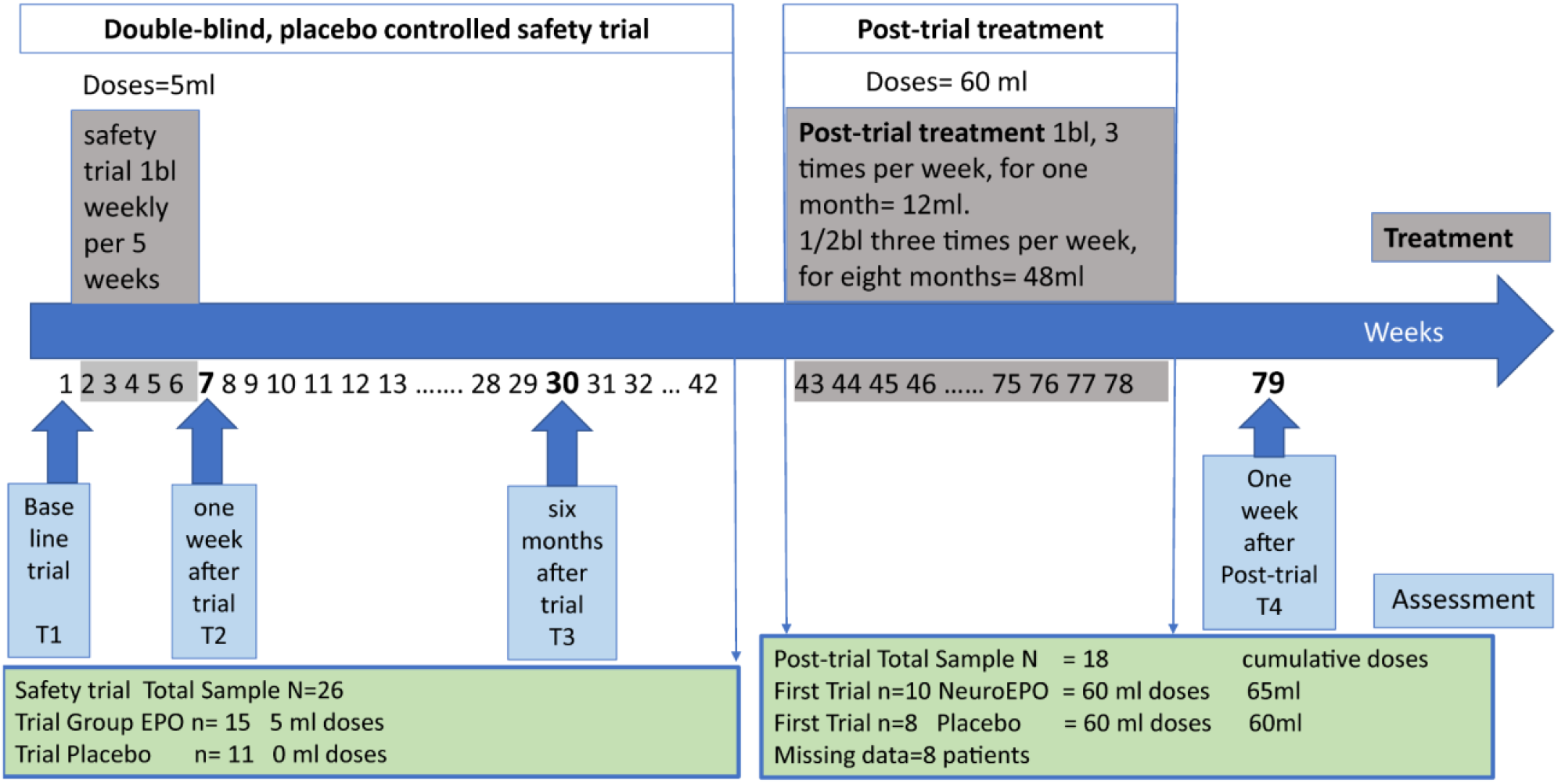
Time of evaluation in weeks (dark blue). The intervention in grey and evaluation time points (T1, T2, T3, and T4) in light blue.

This study was approved by the ethics committee of two institutions: the International Center for Neurological Restoration (CIREN) and the Center for Molecular Immunology (CIM), Cuba, following all the guidelines for clinical trials of the Ministry of Public Health of the Republic of Cuba. All the patients and caregivers received a detailed explanation about the nature and consequences of the study and signed the required informed consent.

### Neuropsychological assessment

Cognitive performance was assessed with a comprehensive neuropsychological battery including global cognitive screening: the Mini-Mental State Examination ^23^and the Dementia Rating Scale ^24^; memory: the Rey Auditory Verbal Learning test (episodic verbal memory), the subtest letter-number sequencing of the Working Memory Index of WAIS III^25^ (Spanish version The Manual Moderno https://www.worldcat.org/title/wais-iii-escala-weschler-de-inteligencia-para-adultos-iii/oclc/54053545) and the Rey Complex Figure, copy and delayed recall/ reproduction of the copy (visuospatial ability and non-verbal memory); executive function: Delis-Kaplan verbal fluency^26^, Trail-Making ^27^ the Stroop color-word Interference test ^28^ and the Frontal Assessment Battery ^29^. Each test was administered at the baseline, one week and six months after the first trial and after the post-trial intervention. The tests were scored according to standard procedures. The patients remained on their usual anti-parkinsonism medication (levodopa and dopaminergic agonists) during the neuropsychological assessments.

### Motor assessment

To assess the motor symptoms, the Unified Parkinson Disease Rating Scale (UPDRS) motor section^30^ was employed. The scale was applied in two conditions: “off” and “on” medication. The analysis of the individual items and total score of the UPDRS scale was evaluated within and between groups at all the timepoints of the trial using item-response theory and t-tests, respectively, for another publication in progress.

### Statistical Analysis

#### Linear Mixed Effect Model

To know the effects of the drug (Dose) on cognitive performance, we selected a multivariate linear mixed-effects model in a longitudinal time-dependent repeated data approach. This is defined for each subject *i* in a sample of *N* subjects, considering the dependent variable of interest Y (cognitive performance) a vector of ni=4 repeated measures as follows

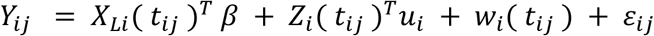

where *X*_*Li*_(*t*_*ij*_) and *Z*_*i*_(*t*_*ij*_) are two vectors of covariates at time *t*_*ij*_ of respective length *p* and *q*. The vector *X*_*Li*_(*t*_*ij*_) is associated with the vector of fixed effects *β*. The vector *Z*_*i*_(*t*_*ij*_), which typically includes functions of time *t*_*ij*_, is associated with the vector of random effects *u*_*i*_.

The vector *u*_*i*_ of q random effects has a zero-mean multivariate normal distribution with variance-covariance matrix *B*. The measurement errors *ε*_*ij*_ are independent Gaussian errors with variance 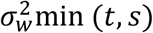.

The dependent variable of interest Y (cognitive performance) is assessed repeatedly in the four timepoints, to test if is a predictor of the drug’s effectiveness.

#### Latent variables

This model uses a latent variable instead of the discrete neuropsychological scores ^31^, because our interest is not the analysis of specific tests individually, but the underlying concept that observable markers measure the cognitive status, the latent phenomena ^32^.

The latent process *𝛬*_*i*_(*t*) is defined in continuous time according to a standard linear mixed model without error of measurement:

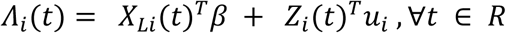

The relationship between the latent process*𝛬*_*i*_(*t*_*ij*_) and the observed value *Y*_*ij*_ at the measurement time *t*_*ij*_ is defined using a linear link function 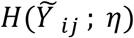. For our model we use a linear function as link for each covariable:

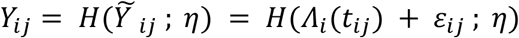

For the statistical analysis, we selected from the comprehensive neuropsychological battery a subset of tests with relative independence and not correlated. We did not select tests and their subtests; and reduced the number of parameters to 6 for a better fit, according to sample size. We included representative measures in each cognitive domain. Global screening (MMSE and DRS total), Memory (RAVLT (Recognition), Figure Rey recall (Memory), Working memory (sequence letter-number), and executive function: (FAB total).

The neuropsychological measurements were collected 4 times (T1 baseline, T2 one week, T3 six months after safety trial) for the N=26 patients and T4 one week after the post-trial for 18 patients. For that reason, some variables have missing values, but the *lcmm* package systematically removed them.

We employed the package *lcmm* in R described by Proust-Lima, (2015)^33^ https://cran.r-project.org/web/packages/lcmm/index.html, specifically the function *multlcmm* to fit latent class mixed models for multivariate data with a common underlying process, in this case, the cognitive performance (observations from the neuropsychological assessment).

#### Covariates

We included several covariates such as gender, age, education, PD progression (duration in years), and severity (Hoehn & Yahr stage). All the covariates were tested in the model.

#### The fixed effect were

– education: in grades (primary 6, secondary 9, high school 12, university 17)
– initage: the initial age in years when the patient was recruited.
– Dose: the cumulative Dose of NeuroEPO received by the patients in each evaluation. Note that we did not consider the main effect “group” because in the post-trial all the patients received the drug without distinction.

This binary condition makes difficult the continuity effect. For that reason, we include the logarithm of the Dose (log(Dose + 1)) to study its impact.

#### The random effects were

– time (timepoints) for the assessment of the cognitive performance of each individual according to the Dose received in each moment.
– subject (their identification number) to control the variability of the repeated measures.

We tested the model with one latent variable (“ng” in the model) to know which model explains better the variability of the markers (neuropsychological tests).

The Wald test statistic was used to test the fixed effects and the goodness-of-fit we employed the maximum log-likelihood, the Akaike Information Criteria (AIC), and the Bayesian Information Criteria (BIC). The models with the minimum AIC and BIC provide the best fit.

## Results

### Participants

The table 1 included the demographic, clinical, cognitive and motor characteristics of the study participants before and after the six-month intervention. Neither age (*p*=0.16), duration of illness (*p*=0.98), familial antecedents of PD (p=0.5), or stage of illness (severity of PD) (p=0.26) were significantly different between the two groups. The comparison between baseline and 6-months after intervention in both groups didn’t show significant differences in the motor variables and Levodopa treatment (UPDRS “on” (p=0.71) and “off” (p=0.88), and LED (p=0.77)). The neuropsychological performance at six months was partially published^18^. We didn’t compare the original sample with the post-trial sub-sample because the division between groups disappeared.

### Cognitive performance

The first step in this analysis was to design a general latent class mixed model fitted by maximum likelihood with one latent class to explain the variability of the dataset. An exploratory model (**AIC=2185 BIC:2215**) found that covariates sex (p=0.43), severity (p=0.35), and progression of the disease (p=0.62) weren’t significant and, for that reason, removed from the final model. See the formula in R notation: mod <-multlcmm(fixed= DRS + FAB + MMSE + sequency + Memory + Recognition ∼ log(Dose+1) + initage + education, random =∼ time, subject = ‘ID’, ng = 1, data = tabla)

The Maximum Likelihood Estimates are included in table 2 below.

**Table 2.** Fixed effects in the longitudinal model.

| **Table 2. Fixed effects in the longitudinal model** |  |  |  |  |
| --- | --- | --- | --- | --- |
|  | Coef. | Se | Wald | p-value |
| intercept (not estimated) | 0.00 |  |  |  |
| Log(Dose+1) | 0.27 | 0.07 | 4.16 | 0.00003 |
| Initage | -0.79 | 0.29 | -2.73 | 0.0063 |
| Education | 0.16 | 0.05 | 2.94 | 0.0032 |

This Model had a good fit **(AIC: 2185.5 BIC: 2215.7**) which can be observed with a residual analysis (see supplementary material).

The model showed a significant linear relation of the latent variable (neuropsychological performance) with the NeuroEPO dose (p=0.00003).Education showed a direct and statistically significant main effect (p=0.003) on the latent variable, indicating that cognitive improvement was related to higher education levels and higher doses.

By contrast, the significant negative effect of the patient’s age at the beginning of the trial indicates better results for younger patients (p=0.006).

To evaluate the relative importance of the markers in the latent process, we obtained the percentage of variance explained by the common latent process and the link function, which expresses the individual contribution to the likelihood of a latent process mixed model. In general, the DRS and the sequence explained the higher variance (more than 50% in each moment), and the MMSE explained the less (between 12 to 17%). See detailed variance in the supplementary material.

The estimated link functions between each test and the underlying latent variable are shown in figure 2.

**Figure 2.**
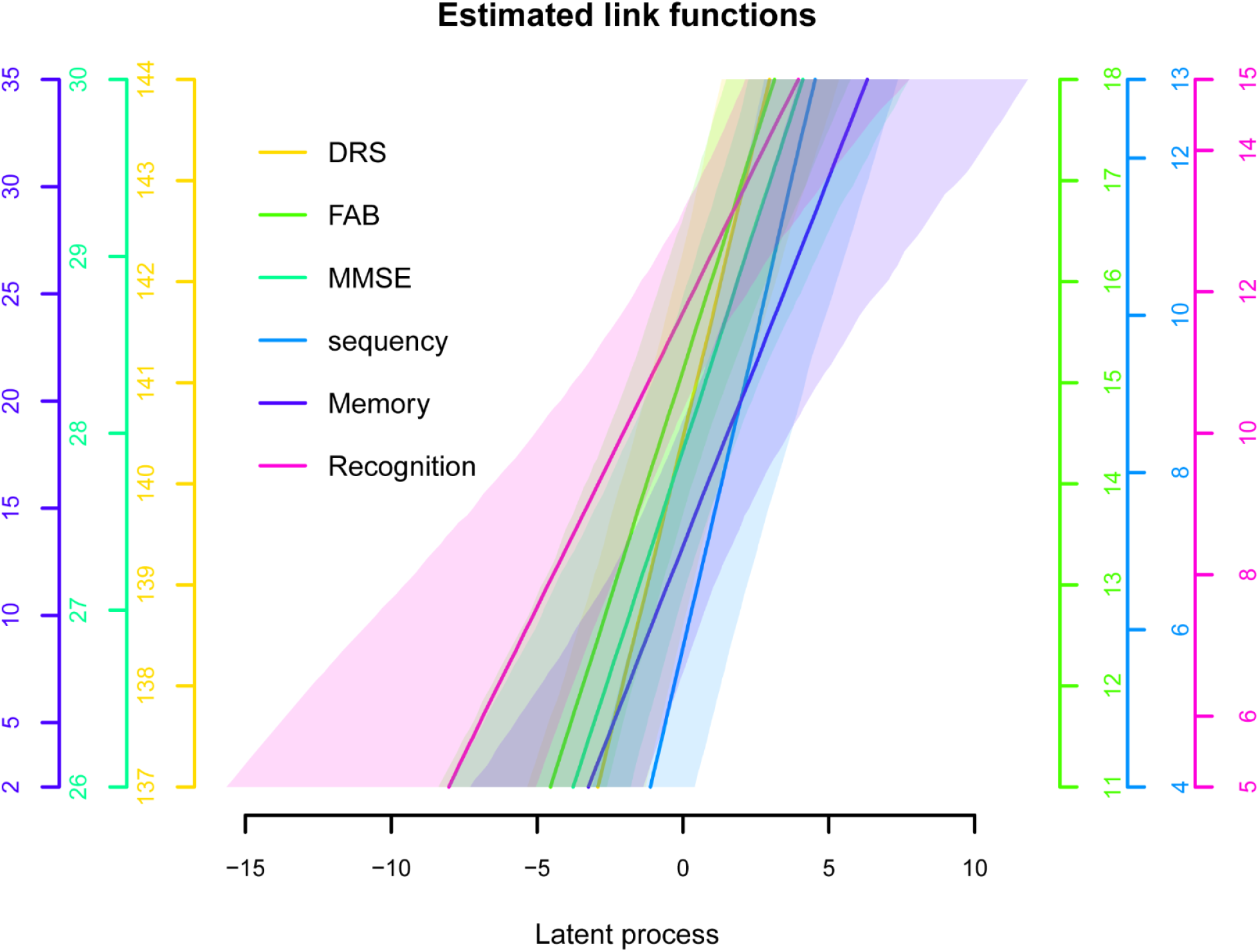
Estimated link functions for the latent variable. The link function of the latent variable with the markers was very significant (p<0.0000).

## Discussion

In this randomized placebo-controlled study, we examined whether there were any changes in cognitive function in a group of (n=15) PD patients treated with a small dose of intranasal NeuroEPO compared to a placebo group (n=11). Later we did a follow-up after 9 months of NeuroEPO treatment when they received an intensive dose of the drug. We found that this formulation of NeuroEPO had a beneficial effect on cognitive performance in all the timepoints evaluated using a longitudinal model.

We selected a linear mixed effect model instead of a t-test or ANOVA to test the cognitive performance between timepoints instead of evaluating each trial separately. Essentially, because a high number of comparisons invalidate any univariate tests (p fishing) and the sample’s small size; we took advantage of the benefits of using a linear mixed effect model to achieve a more robust analysis.

We preferred to use latent variables to explore changes in cognitive performance’s main domains instead of raw neuropsychological scores. The original neuropsychological battery was composed of several subtests which were highly correlated, with different metrics to assess the performance. The multiple directions of the neuropsychological tests assessing cognition and the measurement error associated with the observed variables make it more challenging to interpret the analysis using individual scores. The solution was to look for a common factor underlying the cognitive performance and select a representative cluster of independent tests sufficient to explain them.

Another reason to use the latent variables is to reduce the ceiling-floor effect of global screening tools such as the MMSE and the DRS, which were included, first, because MMSE is the most well-known psychometric test used to describe cognitive aging and second because the Movement Disorder Society has recommended the DRS to study cognition in PD ^34^.

Our preliminary report found^18^ cognitive improvements in several tests (FAB, DRS, Rey figure copy and recall) in both groups (Placebo and NeuroEPO) when comparing baseline with one week and six months after the intervention. In our opinion, this could be explained partially because the first timepoint, only six weeks after the baseline, was influenced by practice and learning effects. And second, because of the placebo effect in PD, mediated through activation of the dopamine system ^35^, and can be elicited by several factors such as the expectation of benefits described in clinical trials of Parkinson’s disease ^36^.

The inclusion in the model of the 4 timepoints across the two trials in the longitudinal study in a latent mixed model allows for balancing all these factors.

Our results highlighted the positive and direct significance of the doses of NeuroEPO (p=0.00003). Note that the doses employed in the double-blind first trial were modest because the nature of the safety trial obliged us to use small, not therapeutic, doses. This disadvantage may have influenced the expression of the cognitive improvement with NeuroEPO relative to Placebo. But could be compensated by including the post-trial with higher and therapeutic doses.

Our results highlighted the protective effect of “education”, and its influence on the NeuroEPO doses, indicating that patients with high educational levels have a better cognitive prognosis. The effect of the age at entry to the trial was significant in a negative direction, showing that the younger the patients were when they started this treatment, the better cognitive outcomes obtained. Many factors, including age, heavily influence the progression of symptoms in PD and the associated cognitive impairment; in that sense, this result is relevant for the clinicians in the recruitment of patients for new protocols. Other covariates such as sex, years of disease progression, and severity didn’t show statistical significance in this trial.

To our knowledge, this is the first study about the cognitive improvement linked to NeuroEPO treatment in PD patients. We can only mention experimental models about the underlying mechanisms that provoke this improvement. Garzon (2018) confirmed in vitro neuroEPO’s protective effect against neuronal damage induced by excitotoxicity, improving antioxidant activity in the neuron, and protecting it from oxidative stress^17^ and showed how NeuroEPO protects cortical neurons from glutamate-induced apoptosis.^37^ In an APPSwe transgenic mice model of Alzheimer’s disease, using a low dose of NeuroEPO, cognitive improvement was observed in behavioral outcomes (spontaneous alternation, place learning in the water-maze, and novel object recognition). A post-mortem analysis of the hippocampus or cortex of the animals showed a decrement in synaptic markers of oxidative stress, neuroinflammation, trophic factors, and beta-amyloid load.^16^

Our findings, even if preliminary, are clinically relevant, given the importance and impact of cognitive deficits on quality of life in PD^5,^ and to date, no treatments have been approved as neuroprotective agents in Parkinson’s Disease.

One limitation is that this study did not include neuroimaging techniques, only behavioral performance. Future studies should adopt a multimodal approach to clarify the neural mechanisms explaining this cognitive improvement.

## Data Availability

All data produced are available online at https://doi.org/10.17605/OSF.IO/M8SJP

## Acknowledgments

The National Natural Science Foundation of China (NSFC) project 61871105 and the CNS Program of UESTC Y0301902610100201 supported the participation of MLB, PAVS, LS, FAR and MJ. The Cuban side was supported by the Ministry of Public Health and Ministry of Science and Technology of the Republic of Cuba. We are grateful to the patients of the International Center for Neurological Restoration, CIREN in La Habana, Cuba, and the staff who volunteered to be part of this study.

## Author Contributions

IPI and MGL conducted the clinical trial, recruited the PD patients, and did all the clinical and motor assessments. ECF was in charge of the neuropsychological assessment. DAG, TRO, IST, and LPR designed the NeuroEPO intervention, doses usage, and provided the drug. LS curated the data and statistical analysis with YRL AMR, FAR, MLB, and PVS. MLB, MJ, and PVS wrote the final version of the manuscript. All authors reviewed the manuscript. MLB, LS, YRL and AMR have equal contribution. PVS and IPI are co-corresponding authors of this paper.

## Availability of data and material

The neuropsychological assessment with clinical and demographic variables are included in the dataset **NeuroEPOtablePD.xls** are available in the OSF repository https://doi.org/10.17605/OSF.IO/M8SJP

